# Drivers of acceptance of COVID-19 proximity tracing apps in Switzerland

**DOI:** 10.1101/2020.08.29.20184382

**Authors:** Viktor von Wyl, Marc Höglinger, Chloé Sieber, Marco Kaufmann, André Moser, Miquel Serra-Burriel, Tala Ballouz, Dominik Menges, Anja Frei, Milo A. Puhan

## Abstract

**Background:** Digital proximity tracing (DPT) apps have been released to mitigate SARS-CoV-2 transmission. But it remains unclear how their acceptance and uptake can be improved. The aim of this study was to investigate SwissCovid app coverage and reasons for not using the app in Switzerland during a time of increasing SARS-CoV-2 incidence.

**Methods:** By use of data collected between 28.09.2020 to 08.10.2020 for a nationwide online panel survey (Covid-19 Social Monitor, n=1’511 participants), socio-demographic and behavioral factors associated with app usage were examined using multivariable logistic regression. Reasons for app non-use were analyzed descriptively.

**Results:** Overall, 46.5% of participants reported using the SwissCovid app (up from 43.9% in a study wave conducted in July 2020).

A higher monthly household income (e.g. OR 1.92 [1.40-2.64] for an income >CHF 10’000 vs. an income ≤ CHF 6’000), more frequent internet use (e.g., daily (reference) vs. less than weekly OR 0.37 [0.16-0.85]), better adherence to mask-wearing recommendations (e.g., always or most of the time (reference) vs. rarely or never OR 0.28 [0.15-0.52]), and being a non-smoker (OR 1.32 [1.01-1.71]) were associated with an increased likelihood for app uptake. Citizenship status (e.g., non-Swiss citizenship 0.61 [0.43-0.87] vs. Swiss citizenship only), and language region (French 0.61 [0.46-0.80], vs. Swiss German) were associated with a lower app uptake probability.

In a randomly selected subsample (n=711) with more detailed information, higher levels of trust in government and health authorities were additionally associated with a higher app uptake probability (e.g., OR 3.13 [1.58-6.22] for high vs. low trust (reference)).

The most frequent reasons for app non-use was lack of perceived benefit of the app (36.8%), 22.8% reported to have no compatible phone, and 22.4% had privacy concerns.

**Conclusion:** Removing technical hurdles and communicating the benefits of DPT-apps are crucial to promote further uptake, compliance, and ultimately to enhance effectiveness of DPT-apps for pandemic mitigation.

## Background

Since safe and effective vaccines against SARS-CoV-2 are currently unavailable, global and national health authorities still rely on non-pharmaceutical interventions in their fight against the ongoing SARS-CoV-2 pandemic. Cornerstones of pandemic mitigation measures include testing, tracing, isolation, and quarantine (TTIQ).(3) Digital proximity tracing (DPT) apps are expected to further enhance conventional TTIQ measures, in particular classic, interview-based contact tracing. Digital proximity tracing apps are a novel, still largely untested health technology, which anonymously records a person’s proximity contacts, that is, other app users who were within a pre-specified radius for a certain amount of time.(4) In case the app user tests positive for SARS-CoV-2, she/he can notify these proximity contacts in an anonymous manner through the app. Detailed explanations of apps following the decentralized, privacy-preserving proximity tracing (DP-3T) design can be found in Box 1 and elsewhere (4, 5).

The rationales for using digital proximity tracing as pandemic mitigation tools are based on a modelling study which found that digital proximity tracing alone has the ability to stop the pandemic spread of SARS-CoV-2.(6, 7) Classic contact tracing is labor- and time-consuming, and exposed contacts can sometimes only be reached and notified with substantial time lags.(8) By comparison, digital proximity tracing can lead to faster notification and earlier self-quarantine of exposed contacts.(5, 6) In addition, digital proximity tracing has a wider reach than classic contact tracing by also including exposed contacts not known by name to the infected person, such as chance encounters in a public space. However, the modelling study further suggests that these expected effects of digital contact tracing depend on several assumptions. Specifically, a large proportion of the population must use the app (e.g., 60% and more if no other mitigation measures are implemented), turnaround time of test results and digital notification of exposed contacts must be within 1-2 days, and notified contacts should enter self-quarantine immediately.(6, 9)

Digital proximity tracing has been developed and implemented with very limited real-life testing. (3, 10) It currently remains unclear whether and to what extent assumptions stated by the modelling analysis are achievable under real world conditions and whether digital proximity tracing technologies can ultimately have a significant impact on pandemic mitigation.(10, 11) However, emerging data from Switzerland indicates that procedural aspects and user behavior has an influence on procedural performance of digital proximity tracing. (12, 13)

Therefore, the present study intended to investigate and synthesize to what extent some of the conditions for digital proximity tracing functioning, namely broad app uptake, were fulfilled during the first three months after the SwissCovid app release in Switzerland. The analysis addressed three main questions. First, which socio-demographic and health-related factors are associated with SwissCovid app usage? Second, what are the most prominent concerns for non-usage of the SwissCovid app? Third, what is known about compliance of app-users with recommended procedures in case of an app notification indicating proximity contact with a SARS-CoV-2-positive app user? These questions were analyzed using data from a nationwide, online survey panel, complemented by publicly available data.

### Box 1

**How the Swiss digital proximity tracing (SwissCovid) app works**

*The digital privacy-preserving proximity tracing (DP-3T) app architecture has become the basis for national digital proximity tracing apps in several countries (e*.*g*., *Ireland, Italy, Germany) and has gained the support of Apple and Google, who provide application programming interfaces (APIs) to support the app’s functionality*.*(1)*

*The Swiss digital proximity app also follows the DP-3T blueprint and has officially been named the “SwissCovid” app. The SwissCovid app was publicly released on 25*.*06*.*2020*.*(2) Similar to other DP-3T inspired apps, smartphones with the SwissCovid app installed will send and receive Bluetooth Low Energy signals to and from other smartphones with the same app. Ephemeral, non-identifiable keys are exchanged and stored locally on smartphones. As Bluetooth signals weaken with increasing phone distance, the signal attenuation can be employed to determine whether another phone was in close proximity (e*.*g. <1*.*5 meters) and for how long. If one of the app-users tests positive for SARS-CoV-2, this person will be issued an activation code (CovidCode), which can be entered into the app. By doing so, the person releases his/her ephemeral keys, which are then uploaded onto a central server system*.

*Smartphones with DP-3T based apps regularly connect to this central server. The uploaded keys of infected persons are downloaded by all smartphones using the SwissCovid app, and the smartphone owner’s locally stored encounter-history (i*.*e. the list of exchanged keys) will be searched for matches with keys of infected persons. If matches fulfilling the criteria for a close proximity encounter (<1*.*5m over at least 15 minutes) are found, the smartphone owner is notified and advised to call an infoline. Notified persons should enter self-quarantine and get tested for SARS-CoV-2 infection. Therefore, the effect of proximity tracing on pandemic containment is mediated by persons being notified as soon as possible about possible exposure risks and by entering quarantine to break transmission chains (“one step ahead”). The contribution of digital proximity tracing apps is that they can notify contacts faster than would be possible in classic contact tracing, and that warnings can also be extended to persons who are chance encounters and not socially connected to the infected person*.

## Methods

### Data Source

The study was based on survey data from the Swiss Covid-19 Social Monitor project. (14) The Covid-19 Social Monitor is a cohort study of randomly selected participants of an existing online panel population. A weighted sample from the panel, stratified based on age, gender and language region, was used in order to make the sample representative of the Swiss population. Participants receive an invitation every 2-6 weeks to complete a survey on various Covid-19 related topics. The survey started on March 30, 2020 and so far 10 study waves with an average response of 1’500 to 1’700 persons from across Switzerland have been conducted.

The Social Monitor collects information on socio-demographic features, comorbidities, and implementation of preventive measures related to Covid-19. In addition, three standardized questions were introduced to gather information about the usage of the Swiss digital proximity tracing app (Supplementary Table 1). The questions were jointly developed by the study investigators, epidemiologists, and infectious disease experts. The standardized SwissCovid app-related questions were first introduced in wave 8 and subsequently used in waves 9 and 10.

The primary data source for these analyses was wave 10 of the Swiss Covid-19 Social Monitor, which was conducted from September 28 to October 8, 2020 and yielded responses from 1’511 participants. Additional data on media use and trust in government, health authorities, or science, was collected for a randomly selected subsample in wave 10 (n=710 participants). Data from 1’299 participants was available for wave 8 (running from July 13 to 20, 2020) as well as wave 10, which was used for analyzing within-person-changes in SwissCovid app use over time and reasons for app non-use over time. Data from waves 8, 9 (August 17 to 25, 2020) and 10 was used to evaluate user responses to app notifications.

### Context of the Pandemic Situation

The observation period for this study starts from app release on June 25, 2020 to approximately 3 months after app release. By early October, the SwissCovid app was downloaded 2.4 million times, and the number of active users was relatively stable at 1.6 million.(15) Compared with Switzerland’s population size of 8.6 million persons of all age groups (6.6 million in the age groups between 18 to 79 years), the number of active app users correspond to a population coverage of approximately 19% (24.2% among those aged 18 to 79 years).

In hindsight, the time period of this survey marked the starting point for large increases in SARS-CoV-2 incidence. There were 8’114 new SARS-CoV-2 cases (positive PCR-tests) reported during the study period from 28.09.2020 to 08.10.2020 in Switzerland.(15, 16) By contrast, the number of new cases was considerably lower in the preceding 11-day period (n=3’644 from 17.09.2020 to 27.09.2020).(15, 16)

### Ethics statement

For the Covid-19 Social Monitor, the Ethics Committee of the Canton Zurich confirmed that it does not fall under the Swiss Human Research Law (BASEC-Nr. Req-2020-00323). Therefore, informed consent was not needed.

### Statistical analysis

#### Factors associated with app uptake

To study the uptake of the SwissCovid app, users and non-users were compared by age (in 10-year categories), sex, citizenship, language region, frequency of internet use, presence of self-reported comorbidities (respiratory diseases, cardiovascular diseases, stroke, hypertension, diabetes, cancer), application of preventive measures (wearing masks, social distancing), education status, household income, citizenship, and smoking status. Persons reporting to use the app permanently or who turn it off occasionally were considered app users. Those who reported not using the app (either with or without intention to do so later) were considered non-users. Additionally, changes in app usage status between waves 8 and 10 were analyzed descriptively among participants who contributed to both waves. Descriptive analyses were performed by summarizing continuous data as medians [interquartile ranges] and categorical data as percentages.

To investigate factors associated with app usage, multivariable logistic regression models were constructed using the characteristics above-mentioned as variables of interest. Age, sex, and comorbidity status were included as a priori fixed co-variables in all models; the remaining variables, including an a priori defined interaction term for age and sex, were added incrementally and kept if the Akaike Information Coefficient (AIC) decreased by 2 points or more upon variable addition.(17, 18) Further logistic regression analyses on the association between app use and media usage and trust in government or science were performed in the subset of participants for whom this information was available. Results from regression analyses are reported as odds ratio (OR) [95% confidence intervals].

#### Investigations into reasons for app non-use

Reasons for non-use of the SwissCovid app were further explored descriptively (N, %) on the basis of given answer options, as well an open answer field for describing other reasons. The analysis was limited to one primary reason for each participant. Socio-demographic and other characteristics as listed above were compared descriptively across the three most frequent reasons for non-use, as well as a fourth group subsuming all other reasons.

All analyses were performed using Stata version 13 (Stata Corp., College Station TX, USA). Two-sided tests of statistical significance were calculated. The level of statistical significance was set at p<0.05. No adjustments for multiple testing were used.

## Results

### Sample characteristics

The wave 10 survey yielded 1’511 responses, and participant characteristics are shown in Table 1. The median age of survey participants was 48 years and 48.8% were females. Almost two thirds (64.5%) were living in the German language region, 22.1% in the French and 13.4% in the Italian language regions. In wave 10, 703 (46.5%) participants reported to have the app installed, 116 out of whom were occasionally switching it off. By comparison, app installation coverage was 43.9% (n=662) in wave 8 (data not shown). Among 1’299 respondents participating in both waves, 75 of 733 (10.2%) app non-users at wave 8 had the app installed by wave 10 (not shown). However, 4.4% of app users in wave 8 have uninstalled the app by wave 10.

**Table 1:** Description of study population, by app use.

|  | **Social Monitor**<br>**(N=1'511)** | **No app use**<br>**(N=808)** | **App use**<br>**(N=703)** |
| --- | --- | --- | --- |
| Age, median [IQR] | 48 [34; 59] | 49 [35; 58] | 46 [34; 59] |
| Female gender | 738 (48.8%) | 389 (48.1%) | 349 (49.6%) |
| <u>Has a partner</u> |  |  |  |
| No partner | 440 (29.1%) | 246 (30.4%) | 194 (27.6%) |
| Living with partner | 951 (62.9%) | 490 (60.6%) | 461 (65.6%) |
| Not living with partner | 120 (7.9%) | 72 (8.9%) | 48 (6.8%) |
| Has children | 163 (10.8%) | 92 (11.4%) | 71 (10.1%) |
| <u>Citizenship</u> |  |  |  |
| Swiss | 1220 (80.7%) | 624 (77.2%) | 596 (84.8%) |
| Swiss and other | 129 (8.5%) | 83 (10.3%) | 46 (6.5%) |
| Non-Swiss | 162 (10.7%) | 101 (12.5%) | 61 (8.7%) |
| <u>Language region</u> |  |  |  |
| German | 975 (64.5%) | 494 (61.1%) | 481 (68.4%) |
| French | 334 (22.1%) | 200 (24.8%) | 134 (19.1%) |
| Ticino | 202 (13.4%) | 114 (14.1%) | 88 (12.5%) |
| <u>Education</u> |  |  |  |
| Only mandatory schooling | 93 (6.2%) | 60 (7.4%) | 33 (4.7%) |
| Completed professional education | 728 (48.2%) | 406 (50.2%) | 322 (45.8%) |
| University, university of applied sciences | 690 (45.7%) | 342 (42.3%) | 348 (49.5%) |
| Currently working | 1066 (70.5%) | 563 (69.7%) | 503 (71.6%) |
| <u>Monthly household income</u> |  |  |  |
| ≤CHF 6000 | 397 (26.3%) | 246 (30.4%) | 151 (21.5%) |
| CHF 6000 - CHF 10000 | 491 (32.5%) | 261 (32.3%) | 230 (32.7%) |
| >CHF 10000 | 343 (22.7%) | 146 (18.1%) | 197 (28.0%) |
| No answer | 280 (18.5%) | 155 (19.2%) | 125 (17.8%) |
| Smoker | 313 (20.7%) | 188 (23.3%) | 125 (17.8%) |
| Self-reported chronic illness** | 378 (25.0%) | 197 (24.4%) | 181 (25.7%) |
| <u>Use of protective masks</u> |  |  |  |
| Always or most of the time | 962 (63.7%) | 494 (61.1%) | 468 (66.6%) |
| Sometimes | 484 (32.0%) | 264 (32.7%) | 220 (31.3%) |
| Rarely or never | 65 (4.3%) | 50 (6.2%) | 15 (2.1%) |
| <u>Staying at home except for essential tasks</u> |  |  |  |
| Always or most of the time | 409 (27.1%) | 224 (27.7%) | 185 (26.3%) |
| Sometimes | 622 (41.2%) | 316 (39.1%) | 306 (43.5%) |
| Rarely or never | 480 (31.8%) | 268 (33.2%) | 212 (30.2%) |
| <u>Frequency of internet use</u> |  |  |  |
| Once daily or several times a day | 1329 (88.0%) | 685 (84.8%) | 644 (91.6%) |
| Once weekly or several days per week | 150 (9.9%) | 99 (12.3%) | 51 (7.3%) |
| Never or less than once weekly | 32 (2.1%) | 24 (3.0%) | 8 (1.1%) |
| <u>Trust in government *</u> |  |  |  |
| Little | 60/712 (8.4%) | 47/375 (12.5%) | 13/337 (3.9%) |
| Somewhat | 163/712 (22.9%) | 102/375 (27.2%) | 61/337 (18.1%) |
| Large | 489/712 (68.7%) | 226/375 (60.3%) | 263/337 (78.0%) |
| <u>Trust in science *</u> |  |  |  |
| Little | 58/710 (8.2%) | 42/374<br>(11.2%) | 16/336 (4.8%) |
| Somewhat | 207/710<br>(29.2%) | 129/374<br>(34.5%) | 78/336<br>(23.2%) |
| Large | 445/710<br>(62.7%) | 203/374<br>(54.3%) | 242/336<br>(72.0%) |
| <u>Questions related to SwissCovid app</u> |  |  |  |
| App user | 587 (38.8%) | `- | `- |
| App user, occasionally switching off the app | 116 (7.7%) | `- | `- |
| Planning to use the app | 53 (3.5%) | `- | `- |
| Has uninstalled app | 66 (4.4%) | `- | `- |
| Not using the app | 689 (45.6%) | `- | `- |

\* Data only available in a randomly selected split-sample including 50% of the full study population.
\*\* Presence of chronic illnesses was defined based on self-report of at least one of the following conditions: asthma, chronic obstructive pulmonary disease (COPD), diabetes, hypertension, cardiovascular disease, stroke, cancer.

### Factors associated with app uptake

Multivariable logistic regression analyses revealed that several factors were associated with app uptake (Table 2, middle column). In the analysis of the full study sample, citizenship status (Swiss and second citizenship OR 0.58 [95% CI 0.40-0.86], non-Swiss citizenship 0.61 [0.43-0.87] vs. Swiss citizenship only), and language region (French-speaking 0.61 [0.46-0.80], Italian-speaking 0.78 [0.57-1.08], vs. German-speaking) were associated with lower app uptake.

**Table 2:** Multivariable logistic regression models for SwissCovid app use as outcome., OR presented

|  | Full sample, N=1'511<br>Univariable OR<br>[95% CI] | Full sample, N=1'511<br>Multivariable OR<br>[95% CI] | Random sub-sample<br>interviewed on trust<br>in government and<br>science, N=712<br>Multivariable OR<br>[95% CI] |
| --- | --- | --- | --- |
| Age (per 10 years) | 1.00 [0.99; 1.01] | 0.99 [0.92; 1.06] | 1.09 [0.98; 1.22] |
| Female Gender (vs. male) | 1.06 [0.87; 1.30] | 1.10 [0.89; 1.36] | 0.94 [0.68; 1.30] |
| <u>Has a partner</u> |  |  |  |
| No partner | Ref. | n.d. | n.d. |
| Living with partner | 1.19 [0.95; 1.50] | n.d. | n.d. |
| Not living with partner | 0.85 [0.56; 1.27] | n.d. | n.d. |
| Has children (vs. not) | 0.87 [0.63; 1.21] | n.d. | n.d. |
| <u>Citizenship</u> | - |  |  |
| Swiss | Ref. | Ref. | Ref. |
| Swiss and other | 0.58 [0.40; 0.85] | 0.58 [0.40; 0.86] | 0.52 [0.28; 0.96] |
| Non-Swiss | 0.63 [0.45; 0.89] | 0.61 [0.43; 0.87] | 0.68 [0.39; 1.20] |
| <u>Language region</u> | - |  |  |
| German | Ref. | Ref. | Ref. |
| French | 0.69 [0.53; 0.89] | 0.61 [0.46; 0.80] | 0.56 [0.37; 0.84] |
| Ticino | 0.79 [0.58; 1.08] | 0.78 [0.57; 1.08] | 0.90 [0.54; 1.51] |
| <u>Education</u> | - |  |  |
| Only mandatory schooling | Ref. | Ref. | Ref. |
| Completed professional education | 1.44 [0.92; 2.26] | 1.32 [0.83; 2.12] | 1.23 [0.57; 2.63] |
| University, university of applied sciences | 1.85 [1.18; 2.90] | 1.50 [0.94; 2.42] | 1.58 [0.73; 3.45] |
| Currently working (vs. not working) | 0.91 [0.73; 1.14] | n.d. | n.d. |
| <u>Monthly household income</u> | - |  |  |
| ≤CHF 6000 | Ref. | Ref. | Ref. |
| CHF 6000 - CHF 10000 | 1.44 [1.10; 1.88] | 1.29 [0.97; 1.71] | 1.14 [0.75; 1.74] |
| >CHF 10000 | 2.20 [1.64; 2.95] | 1.92 [1.40; 2.64] | 1.53 [0.94; 2.48] |
| No answer | 1.31 [0.96; 1.79] | 1.18 [0.85; 1.63] | 1.06 [0.66; 1.71] |
| Being non-smoker (vs. smoker) | 1.40 [1.09; 1.81] | 1.32 [1.01; 1.71] | 1.51 [1.02; 2.25] |
| Self-reported chronic illness** (vs. none) | 1.08 [0.85; 1.36] | 1.11 [0.87; 1.43] | 0.88 [0.61; 1.27] |
| <u>Use of protective masks</u> | - |  |  |
| Always or most of the time | Ref. | Ref. | Ref. |
| Sometimes | 0.88 [0.71; 1.10] | 0.75 [0.60; 0.96] | 0.77 [0.54; 1.10] |
| Rarely or never | 0.32 [0.18; 0.57] | 0.28 [0.15; 0.52] | 0.32 [0.12; 0.86] |
| <u>Staying at home except for essential tasks</u> |  |  |  |
| Always or most of the time | Ref. | n.d. | n.d. |
| Sometimes | 1.17 [0.91; 1.51] | n.d. | n.d. |
| Rarely or never | 0.96 [0.73; 1.25] | n.d. | n.d. |
| <u>Frequency of internet use</u> | - |  |  |
| Once daily or several times a day | Ref. | Ref. | Ref. |
| Once weekly or several days per week | 0.55 [0.38; 0.78] | 0.55 [0.38; 0.80] | 0.59 [0.35; 1.00] |
| Never or less than once weekly | 0.35 [0.16; 0.79] | 0.37 [0.16; 0.85] | 0.32 [0.11; 0.95] |
| <u>Trust in Government / Health Authorities*</u> | - |  |  |
| Little | Ref. | n.a. | Ref. |
| Somewhat | 2.16 [1.08; 4.32] | n.a. | 1.71 [0.82; 3.58] |
| Large | 4.21 [2.22; 7.97] | n.a. | 3.13 [1.58; 6.22] |
| <u>Trust in science*</u> |  |  |  |
| Little | Ref. | n.a. | n.d. |
| Somewhat | 1.59 [0.84; 3.01] | n.a. | n.d. |
| Large | 3.13 [1.71; 5.73] | n.a. | n.d. |

Abbreviations: CI Confidence interval, OR Odds ratio, n.a., not available; n.d. not done/not included because not improving model fit; Ref., Reference.
\* Data only available in a randomly selected split-sample including 50% of the full study population.
\*\* Presence of chronic illnesses was defined based on self-report of at least one of the following conditions: asthma, chronic obstructive pulmonary disease (COPD), diabetes, hypertension, cardiovascular disease, stroke, cancer.

By contrast, a higher monthly household income (OR 1.92 [1.40-2.64] for an income >CHF 10’000 vs. an income ≤ CHF 6’000; 1 CHF equals 0.93 EUR or 1.10 US$), more frequent internet use (daily OR 1 (reference) vs. less than weekly OR 0.37 [0.16-0.85]), better adherence to mask wearing recommendations (always or most of time OR 1 (reference) vs. rarely or never OR 0.28 [0.15-0.52]), and being a non-smoker (OR 1.32 [1.01-1.71]) were associated with increased app uptake.

The same model was also applied to the random sub-sample (Table 2, last column), which provided additional information on trust in government and science (n=712). Of note, the odds ratios of variables included in both multivariable models (full and sub-sample) were not altered substantially, but confidence intervals became wider due to the lower sample size. Furthermore, increasing levels of trust in government and health authorities was also associated with a higher app uptake probability (OR 3.13 [1.58-6.22] for high vs. low trust (reference)), whereas the inclusion of trust in science did not improve the multivariable model fit.

### Reasons for non-use of app

The responses of persons who reported not to use the app were analyzed further with respect to reasons for non-use (Table 3). This group included both persons who stated that they are planning to use the app and those who do not plan to use it. Overall, the most important reasons for not installing the app were a *perceived lack of usefulness of the app* (36.8%), followed by *not having a suitable smartphone or operating system* (22.8%), and *concerns about privacy* (22.4%). Other reasons amounted to 18.0% and included *not knowing the app, doubts about technological reliability, concerns about excessive battery usage*, among others.

**Table 3:** Reasons for non-use of the SwissCovid app

|  | May install app later | App not installed | Uninstalled app | All |
| --- | --- | --- | --- | --- |
| N | 53 (100%) | 689 (100%) | 66 (100%) | 808 (100%) |
| Perceived as not useful | 15 (28.3%) | 262 (38.0%) | 20 (30.3%) | 297 (36.8%) |
| Not the right phone | 18 (34.0%) | 158 (22.9%) | 8 (12.1%) | 184 (22.8%) |
| Concerned about privacy | 8 (15.1%) | 164 (23.8%) | 9 (13.6%) | 181 (22.4%) |
| Don't know the app | 2 (3.8%) | 25 (3.6%) | 0 | 27 (3.3%) |
| Technical doubts about reliability, maturity | 1 (1.9%) | 20 (2.9%) | 4 (6.1%) | 25 (3.1%) |
| Concerned about battery use | 1 (1.9%) | 8 (1.2%) | 11 (16.7%) | 20 (2.5%) |
| Don't believe in seriousness of Corona; lack of trust in government | 0 | 9 (1.3%) | 1 (1.5%) | 10 (1.2%) |
| Inertia, not had the time yet | 5 (9.4%) | 2 (0.3%) | 0 | 7 (0.9%) |
| Opposed out of principle, no specific reason | 0 | 7 (1%) | 0 | 7 (0.9%) |
| Don't want Bluetooth permanently on | 0 | 3 (0.4%) | 2 (3.0%) | 5 (0.6%) |
| Worried about consequences/quarantine | 0 | 2 (0.3%) | 2 (3.0%) | 4 (0.5%) |
| Currently outside of Switzerland | 1 (1.9%) | 3 (0.4%) | 0 | 4 (0.5%) |
| Would have to turn off app at work | 0 | 2 (0.3%) | 1 (1.5%) | 3 (0.4%) |
| Would feel stressed/scared by app use | 0 | 2 (0.3%) | 0 | 2 (0.2%) |
| Already protecting themselves, rarely leave the house | 0 | 1 (0.1%) | 0 | 1 (0.1%) |

When compared to responses from wave 8, the percentage of persons reporting a perceived lack of usefulness (27.0%) was considerably lower, whereas differences for the other two reasons were less pronounced (not having the right phone 26.1%, privacy concerns 23.9%, and other reasons 23.0%; data not shown).

The distribution of reasons for non-use also varied across levels of intention for using the app later (maybe, no, or already uninstalled the app). While lack of perceived benefits was the dominant reason for not installing (38.0%) and for having uninstalled (30.3%) the app, 34.0% reported not to have a compatible phone among persons with an intent to install the app at a later time point. Further noteworthy, excessive battery consumption also appeared to be an important reason for uninstalling the app (16.7%).

The descriptive comparison of socio-demographic and other characteristics across the three major reasons for app non-use (and a fourth category subsuming all other reasons, Table 4) suggests that some reasons may be more prevalent in specific subgroups. The sub-population stating problems with installing the app (“not the right phone”) was the oldest (median age 57.5 years), had the highest burden of chronic comorbidities (33.2%), but tended to have high trust in government (large trust category: 77.5%) and science (large trust category: 68.5%) compared with the other subgroups. By contrast, those reporting privacy concerns for non-use were younger (median age 44 years), were more frequently living in the French-speaking part of Switzerland (35.9%), and generally had less trust in the government (large trust category: 43.8%) or science (large trust category: 37.5%).

**Table 4:** Socio-demographic characteristics of persons not using the app, stratified by reason for non-use (three most frequent and other)

|  | Not right phone<br>(N=184) | Privacy concerns<br>(N=181) | Not useful<br>(N=297) | Other reason<br>(N=146) | p-value |
| --- | --- | --- | --- | --- | --- |
| Age, median [IQR] | 57.5 [44.5; 67] | 44 [35; 54] | 46 [31; 57] | 44 [31; 57] | <0.0001 |
| Female gender | 95 (51.6%) | 102 (56.4%) | 120 (40.4%) | 72 (49.3%) | 0.005 |
| <u>Has a partner</u> |  |  |  |  | 0.511 |
| No partner | 45 (24.5%) | 58 (32.0%) | 100 (33.7%) | 43 (29.5%) |  |
| Living with partner | 123 (66.8%) | 106 (58.6%) | 172 (57.9%) | 89 (61.0%) |  |
| Not living with partner | 16 (8.7%) | 17 (9.4%) | 25 (8.4%) | 14 (9.6%) |  |
| Has children | 15 (8.2%) | 21 (11.6%) | 27 (9.1%) | 29 (19.9%) | 0.003 |
| <u>Citizenship</u> |  |  |  |  | 0.129 |
| Swiss | 152 (82.6%) | 132 (72.9%) | 232 (78.1%) | 108 (74.0%) |  |
| Swiss and other | 10 (5.4%) | 21 (11.6%) | 35 (11.8%) | 17 (11.6%) |  |
| Non-Swiss | 22 (12.0%) | 28 (15.5%) | 30 (10.1%) | 21 (14.4%) |  |
| <u>Language region</u> |  |  |  |  | 0.006 |
| German | 115 (62.5%) | 100 (55.2%) | 188 (63.3%) | 91 (62.3%) |  |
| French | 41 (22.3%) | 65 (35.9%) | 64 (21.5%) | 30 (20.5%) |  |
| Ticino | 28 (15.2%) | 16 (8.8%) | 45 (15.2%) | 25 (17.1%) |  |
| <u>Education</u> |  |  |  |  | 0.514 |
| Only mandatory schooling | 16 (8.7%) | 16 (8.8%) | 22 (7.4%) | 6 (4.1%) |  |
| Completed professional education | 90 (48.9%) | 83 (45.9%) | 157 (52.9%) | 76 (52.1%) |  |
| University, university of applied sciences | 78 (42.4%) | 82 (45.3%) | 118 (39.7%) | 64 (43.8%) |  |
| Currently working | 97 (52.7%) | 140 (77.3%) | 221 (74.4%) | 105 (71.9%) | <0.0001 |
| <u>Monthly household income</u> |  |  |  |  | 0.027 |
| ≤CHF 6000 | 65 (35.3%) | 52 (28.7%) | 87 (29.3%) | 42 (28.8%) | 0.027 |
| CHF 6000 - CHF 10000 | 62 (33.7%) | 48 (26.5%) | 104 (35.0%) | 47 (32.2%) |  |
| >CHF 10000 | 25 (13.6%) | 30 (16.6%) | 61 (20.5%) | 30 (20.5%) |  |
| No answer | 32 (17.4%) | 51 (28.2%) | 45 (15.2%) | 27 (18.5%) |  |
| Smoker | 36 (19.6%) | 47 (26.0%) | 78 (26.3%) | 27 (18.5%) | 0.138 |
| Self-reported chronic illness** | 61 (33.2%) | 43 (23.8%) | 66 (22.2%) | 27 (18.5%) | 0.011 |
| <u>Use of protective masks</u> |  |  |  |  | 0.077 |
| Always or most of the time | 127 (69.0%) | 110 (60.8%) | 172 (57.9%) | 85 (58.2%) |  |
| Sometimes | 50 (27.2%) | 62 (34.3%) | 106 (35.7%) | 46 (31.5%) |  |
| Rarely or never | 7 (3.8%) | 9 (5.0%) | 19 (6.4%) | 15 (10.3%) |  |
| <u>Staying at home except for essential tasks</u> |  |  |  |  | 0.011 |
| Always or most of the time | 54 (29.3%) | 48 (26.5%) | 83 (27.9%) | 39 (26.7%) |  |
| Sometimes | 90 (48.9%) | 69 (38.1%) | 108 (36.4%) | 49 (33.6%) |  |
| Rarely or never | 40 (21.7%) | 64 (35.4%) | 106 (35.7%) | 58 (39.7%) |  |
| <u>Frequency of internet use</u> |  |  |  |  | 0.014 |
| Once daily or several times a day | 142 (77.2%) | 160 (88.4%) | 257 (86.5%) | 126 (86.3%) |  |
| Once weekly or several days per week | 32 (17.4%) | 14 (7.7%) | 36 (12.1%) | 17 (11.6%) |  |
| Never or less than once weekly | 10 (5.4%) | 7 (3.9%) | 4 (1.3%) | 3 (2.1%) |  |
| <u>Trust in government *</u> |  |  |  |  | 0.001 |
| Little | 5 (5.6%) | 16 (20.0%) | 12 (8.6%) | 14 (21.2%) |  |
| Somewhat | 15 (16.9%) | 29 (36.3%) | 43 (30.7%) | 15 (22.7%) |  |
| Large | 69 (77.5%) | 35 (43.8%) | 85 (60.7%) | 37 (56.1%) |  |
| <u>Trust in science *</u> |  |  |  |  | 0.004 |
| Little | 7 (7.9%) | 13 (16.3%) | 12 (8.6%) | 10 (15.2%) |  |
| Somewhat | 21 (23.6%) | 37 (46.3%) | 47 (33.8%) | 24 (36.4%) |  |
| Large | 61 (68.5%) | 30 (37.5%) | 80 (57.6%) | 32 (48.5%) |  |

\* Data only available in a randomly selected split-sample including 50% of the full study population (n=712).
\*\* Presence of chronic illnesses was defined based on self-report of at least one of the following conditions: asthma, chronic obstructive pulmonary disease (COPD), diabetes, hypertension, cardiovascular disease, stroke, cancer.

Demographics of subpopulations reporting the remaining two reasons (“not useful”, “other reasons”) did not reveal specific patterns.

### SwissCovid app notifications and user response

In three survey waves (8, 9, and 10), a total of 15 persons reported to have received an app notification: 2 persons in wave 8 (July), 6 persons in wave 9 (August), and 7 persons in wave 10 (October). Overall, 8 of those 15 (53.3%) persons reported to have called the recommended infoline, 7 reported not to have undertaken any steps, and 1 person undertook other steps, which were left unspecified.

Since wave 10, participants are also asked about whether they have undergone SARS-CoV-2 testing in the past 4 weeks and what their test results were. Of 5 persons calling the infoline at wave 10, 2 (40.0%) persons reported to have gotten tested for SARS-CoV-2, and 1 (20.0%) person reported to have been tested positive.

## Discussion

By analyzing information on the use of the SwissCovid app from a longitudinal online panel, we looked at factors related to the use of the digital proximity tracing app in Switzerland.

Our data suggest that, three months after app release, 46.5% of the survey respondents had downloaded the app (of whom 38.8% had the SwissCovid app permanently activated). This percentage is an overestimation of actual app coverage in the general population and most likely caused by the above-average affinity for such technologies of online panel participants. Also, social desirability might have led to some over-reporting of app usage, despite the survey being an anonymous online survey. (19) In early October 2020, the official number of active app users was estimated at 1.6 Mio.(15) This number implies that around 1 in 4 (24.2%) adults residing in Switzerland were actively using the app. A recent modelling study suggests that this uptake proportion may in fact be sufficient to reduce the number of new infections to “manageable levels”.(20).

We also deduced a number of population characteristics that may influence the uptake of the SwissCovid app. For example, younger age, higher income, or being a non-smoker were associated with greater app uptake. By contrast, having foreign (non-Swiss) nationality, or living in the French- or Italian-speaking parts of Switzerland were associated with lower uptake. Furthermore, app uptake was associated with the level of trust placed in the government and in health authorities. Moreover, following recommended preventive measures and wearing masks, in particular, were also associated with a higher likelihood to use the app, which could signal higher levels of awareness or worry related to the Covid-19 epidemic or greater health consciousness.

We further investigated participants’ stated reasons for non-use of the SwissCovid app, which were dominated by technical aspects (i.e., not having a suitable smartphone or operating system), privacy concerns, and perceived lack of usefulness. Ignorance or lack of information about the app seemed no relevant reason as only 3% indicated not knowing the app as the reason for their non-use. Privacy concerns as a reason for non-use was associated with a lack of trust in the government and health authorities, as well as with a migration background. By contrast, the group hindered from app use by technical aspects seemed to be more trustful in the government but tended to be older. Therefore, streamlining installation processes and establishing compatibility with older phones may be worthwhile in order to increase uptake in this subgroup.

By contrast, the prevalent privacy concerns and trust issues are harder to tackle. Although the SwissCovid app implements privacy by design, the fact that the app relies on application programming interfaces (APIs) provided by Google and Apple is sometimes criticized. These concerns should be addressed by communication efforts, which, for example, could also focus on personal experiences of app users and tell success stories. The latter may also increase motivations for app use among the substantial fraction (37%) of non-users citing the lack of personal or general benefits of the app as the main reason for their non-use. But there is also evidence that external factors, such as the overall pandemic situation, impacts the perceived benefits. SARS-CoV-2 infections increased rapidly in Switzerland in the second half of October 2020, and the number of active app users also rose by 200’000 persons.(15)

To our knowledge, this is the first study to systematically investigate digital proximity tracing app uptake and the reasons for app non-use in Switzerland. One survey has been conducted in Switzerland since the app release in late June 2020 among 1’000 Swiss individuals.(21) This study, whose data have not been published in detail, yielded that 43% of the Swiss population are using or considering to use the Swiss proximity tracing app, with higher percentages among younger respondents. Our study results show similar proportions of app users, but also shed further light on motivations or barriers for app use. Furthermore, a key strength of our study was the availability of data from different survey waves, which allowed us to verify the robustness of our findings. Furthermore, our sample of 1500 persons is based on a random sample and is therefore likely to be quite representative in various regards for the Swiss population. This is also reflected by the close match of our projected number of app users with official numbers. However, we cannot fully exclude potential biases such as over-reporting or social desirability bias regarding app use. In addition, the fact that the Social Monitor sample was drawn from an online panel population led to an over-estimation of the app usage of the general population.

To summarize, our study yielded a clearer understanding of motivations, barriers and other factors associated with the uptake of digital proximity tracing apps. Our data point to complex interactions between motivations, trust, and incentives. Therefore, communication efforts to promote the app use should convey messages for different subgroups and should particularly focus on successes and beneficial effects of the app.

## Data Availability

The datasets generated during and/or analysed during the current study are available from the corresponding author on reasonable request.

## Acknowledgements

We thank the participants of the the Covid-19 Social Monitor project for their important contribution.

The Social Monitor study has received funding from the Federal Office of Public Health and from Health Promotion Switzerland.

The Funders have no influence on the design, conduct, analyses and publications.

## Author Contributions

VVW conceived and designed the work, conducted the statistical analysis, and drafted the manuscript. C.S., M.S.B., D.M., T.B., M.H., A.M., M.K., A.F. & M.A.P provided input on the analytic strategy. C.S. performed parts of the statistical analysis. M.H., A.M., and M.A.P. designed the Covid-19 Social Monitor and collected the data. M.H. and A.M. prepared the Social Monitor datasets.

All authors contributed to the interpretation of the data and critically revised the manuscript. All the authors gave final approval of the completed manuscript version and are accountable for all aspects of the work.

## Data availability

**Supplementary Table 1:** Standardized questions on SwissCovid app use in the Social Monitor

|  |  |
| --- | --- |
| The SwissCovid App has been launched by the Swiss Federal Office of Public Health to warn smartphone users in case of possible exposure risks. The app records, if a contact has been in close proximity of 1.5m or less for longer than 15 minutes.<br>If an app user tested positive for the Coronavirus, she or he can anonymously notify other app users, who were in close proximity during the infectious period. |  |
|  | Are you using the SwissCovid App?<br>• Yes, permanently<br>• Yes, but sometimes I turn off Bluetooth to pause the SwissCovid App<br>• No, but I am planning to use it<br>• No<br>• Since wave 10: No, I have uninstalled the SwissCovid App |
| Filter If **No** or **No, but...**: | Why are you currently not using the SwissCovid App?<br>• I have not heard about the app<br>• I don't think the app is useful for me<br>• I can't install the app (e.g., owing to technical difficulties or because I do not own an Android or iOS smartphone)<br>• I fear for my privacy and protection of my data<br>• Other reasons, comment field |
| Filter if **yes** or **yes, but**: | Were you ever notified by the SwissCovid App that you have been in close proximity to a Corona-positive person?<br>• No, I have never received a notification<br>• Yes, I called the recommended Infoline SwissCovid<br>• Yes, I undertook other steps; comment field: which?<br>• Yes, but I did not undertake any steps |

